# SARS-CoV-2 vaccine effectiveness and breakthrough infections in maintenance dialysis patients

**DOI:** 10.1101/2021.09.24.21264081

**Authors:** Harold J. Manley, Gideon N. Aweh, Caroline M. Hsu, Daniel E. Weiner, Dana Miskulin, Antonia M. Harford, Doug Johnson, Eduardo K. Lacson

## Abstract

Among patients receiving maintenance dialysis with a national US dialysis provider, fully vaccinated dialysis patients were significantly less likely to be diagnosed with COVID-19 or be hospitalized than unvaccinated patients. Ad26.COV2.S/Janssen vaccine had significantly worse outcomes, and mRNA-1273/Moderna the best. Among the 27 patients with breakthrough COVID-19 and anti-spike IgG antibodies measured, 23/27 (85%) breakthrough COVID-19 cases occurred at titers <2 U/L (lower limit of a positive response); 14 of these patients never developing Ab titers above 2 U/L. Only 3/27 were receiving immunosuppression. The potential use of antibody titers to guide vaccinations should be explored.

---

Maintenance dialysis patients experience significant COVID-19–associated morbidity and mortality.^1,2^ Studies show that 2 doses of the SARS-CoV-2 mRNA vaccines elicit a seroresponse in most (>90%) maintenance dialysis patients, albeit with lower levels than in the general population.^3-6^The impact of this lesser response as well as potentially more rapidly waning immunity on clinical outcomes is not known. Accordingly we describe the incidence of COVID-19 infection and COVID-19 related hospitalization among unvaccinated, partially vaccinated and fully vaccinated dialysis patients from a national dialysis provider within the US between February 1 and August 26, 2021, as well as available SARS-CoV-2 vaccine immunoglobulin G spike antibody (SAb-IgG) titers in fully vaccinated patients with breakthrough infections.

## Methods

All adult maintenance dialysis (home and in-center) patients without known prior COVID-19 infection prior to February 1, 2021 treated at Dialysis Clinic, Inc. (DCI) outpatient dialysis clinics between February 1 and August 26, 2021 were included. Patient demographic and clinical variables were obtained from DCI’s electronic health record database, de-identified and aggregated. This retrospective evaluation was reviewed and approved by WCG IRB (Work Order 1-1456342-1).

Patients were categorized into three cohorts per Centers for Disease Control and Prevention (CDC) criteria: unvaccinated, partially vaccinated (defined as 14 days after the first mRNA vaccine dose up to 13 days after the 2^nd^ mRNA vaccine dose), or fully vaccinated (≥14 days after completing the manufacturer recommended final dose).^4^ Patients could move from being unvaccinated to partially vaccinated to fully vaccinated and contribute days-at-risk for each applicable category as long as they remained COVID-19-free.

Primary study outcomes were new COVID-19 diagnoses and COVID-19 related hospitalization during the study period. Using logistic regression, rates of COVID-19 infection and COVID-19-related hospitalization were compared by vaccine status and vaccine type, adjusting for baseline patient characteristics that differed by vaccination status (Details in Supplemental Methods; **Table S1**). Secondary analyses stratified based on SARS-CoV-2 delta variant dominance in the US (February 1 -June 26, 2021 vs June 27 -August 27, 2021).^7^

In descriptive analyses, breakthrough infections and available anti-spike immunoglobulin G antibody (SAb-IgG) levels were assessed relative to baseline, defined as the date of full immunization. Antibody values range from 1 to 20 U/L, with minimum detectable level of ≥1 U/L per manufacturer, although the DCI Lab validated threshold of ≥2 U/L was used.^3,8^ Statistical analyses were performed using SAS v9.4.

## Results

Among 15,251 maintenance dialysis patients at DCI facilities during the study period, 10,576 (69%) were fully vaccinated (5,905 (56%) with mRNA-1273/Moderna, 4,172 (39%) BNT162b2/Pfizer, 499 (5%) Ad26.COV2.S/Janssen), 671 (4%) were partially vaccinated (380 (57%) mRNA-1273/Moderna, 291 (43%) BNT162b2/Pfizer) and 4,004 (26%) were unvaccinated. Mean age of patients was 63±15 years old, with median 2.30 (IQR 4.59) years receiving on dialysis, 87% treated by hemodialysis, and 58% with diabetes (**Table S1**).

### Vaccination status, COVID-19 cases and outcomes

Among 650 documented COVID-19 cases, 405 (62%) occurred in unvaccinated patients. Among fully vaccinated patients, there were 1.20 COVID-19 cases per 10,000 patient days. In contrast, partially vaccinated patients and unvaccinated patients experienced more cases (2.45 per 10,000 patient-days among partially vaccinated patients [adjusted OR 1.97 (1.50, 2.58)] and 3.35 cases per 10,000 patient-days among unvaccinated patients [adjusted OR 2.56 (2.12, 3.09)], respectively). The incidence of COVID-19 related hospitalization, was 1.03 hospitalizations per 10,000 patient days in unvaccinated and 0.43 per 10,000 patient-days in vaccinated patients [adjusted OR 2.21 (1.61, 3.02)](**Table**). There were 27 COVID-19 related deaths during the study period: 17 (70%) among unvaccinated, 1 (4%) among partially vaccinated, and 9 (33%) among fully vaccinated patients. The incidence rate of COVID-19 related deaths within 30 days of COVID-19 diagnosis among unvaccinated patients was twice that observed in fully vaccinated patients [0.13 vs. 0.06 deaths per 10,000 patient-days; adjusted OR 2.52 (1.09, 5.85)]. In both the pre-COVID-19 delta variant (February 1-June 26, 2021) and COVID-19 delta dominant (June 27-August 27, 2021) periods, the incidence rate ratio for COVID-19 and COVID-19 related hospitalization remained significantly higher in unvaccinated patients when compared to fully vaccinated patients (**Tables S2-3**).

**Table.** New and breakthrough COVID-19 infections and COVID related hospitalizations between February 1 and August 26, 2021

| Status/Category | Person Days-at risk | # Events | Incidence Rates per 10,000 Patient Days | Incidence Rate Ratio (95% Confidence Interval) |  |
| --- | --- | --- | --- | --- | --- |
|  |  |  |  | Unadjusted | Adjusted |
| **COVID Cases by Vaccination Status\*** |  |  |  |  |  |
| **Unvaccinated (N = 4,004)** | 1,210,311 | 405 | 3.35 | 2.79 (2.33, 3.34) | 2.56 (2.12, 3.09) |
| **Partially Vaccinated (N = 671)** | 313,828 | 77 | 2.45 | 2.04 (1.56, 2.68) | 1.97 (1.50, 2.58) |
| **Fully Vaccinated (N = 10,576)** | 1,399,238 | 168 | 1.20 | Reference | Reference |
| **COVID Cases by Vaccine Type** |  |  |  |  |  |
| **Ad26.COV2.S/Janssen (N = 499)** | 61,862 | 72 | 2.42 | 2.65 (1.52, 4.63) | 2.33 (1.33, 4.08) |
| **BNT162b2/Pfizer (N = 4,172)** | 550,322 | 81 | 1.47 | 1.61 (1.17, 2.17) | 1.58 (1.15, 2.17) |
| **mRNA-1273/Moderna (N = 5,905)** | 787,054 | 15 | 0.91 | Reference | Reference |
| **COVID-related Hospitalization by Vaccination Status\*** |  |  |  |  |  |
| **Unvaccinated (N=4,004)** | 1,315,290 | 136 | 1.03 | 2.40 (1.78, 3.24) | 2.21 (1.61, 3.02) |
| **Partially Vaccinated (N=671)** | 335,220 | 32 | 0.95 | 2.22 (1.45, 3.40) | 2.11 (1.38, 3.23) |
| **Fully Vaccinated (N=10,576)** | 1,440,511 | 62 | 0.43 | Reference | Reference |
| **COVID-related Hospitalization by Vaccine Type** |  |  |  |  |  |
| **Ad26.COV2.S/Janssen (N = 499)** | 64,460 | 6 | 0.93 | 3.77 (1.52, 9.40) | 3.26 (1.29, 8.21) |
| **BNT162b2/Pfizer (N = 4,172)** | 565,469 | 36 | 0.64 | 2.58 (1.49, 4.46) | 2.55 (1.47, 4.42) |
| **mRNA-1273/Moderna (N = 5,905)** | 810,582 | 20 | 0.25 | Reference | Reference |
| **COVID-related Death by Vaccination Status\*** |  |  |  |  |  |
| **Unvaccinated**<br>**(N=4,004)** | 1,315,290 | 17 | 0.13 | 2.07 (0.92,4.64) | 2.52 (1.09,5.85) |
| **Partially Vaccinated**<br>**(N=671)** | 335,220 | 1 | 0.03 | 0.48 (0.06,3.77) | 0.48 (0.06,3.75) |
| **Fully Vaccinated**<br>**(N=10,576)** | 1,440,511 | 9 | 0.06 | Ref | Ref |

\* Although patients can contribute time to any vaccination status, the N in the first column refers to patients' status at the end of follow-up. Unvaccinated includes patients who never received a vaccine or recipients of a single dose of a vaccine within 14 days of vaccine receipt; partially vaccinated includes patients who were $\geq 14$ days after the first mRNA vaccine dose but $< 14$ days after the 2nd mRNA vaccine dose; fully vaccinated patients includes all patients $\geq 14$ days after the last vaccine dose.

### Vaccine type and breakthrough COVID-19

There were 168 (26% of total cases) that occurred among those considered fully vaccinated, most (n=142; 85%) during the delta dominant period. The median time from being considered fully vaccinated to COVID-19 diagnosis was 127 [98-151] days, and 36 (21%) met CDC criteria for immunosuppressed. Breakthrough COVID-19 infection and COVID-19 related hospitalizations were more likely with the BNT162b2/Pfizer and Ad26.COV2.S/Janssen vaccines as compared to the mRNA-1273/Moderna vaccine (**Table**), with similar results seen during the pre-delta and the COVID-19 delta dominant variant period (**Tables S2 and S3**).

### SAb-IgG level and breakthrough COVID-19

SAb-IgG levels were available in 27 breakthrough infections (BNT162b2/Pfizer N=10, mRNA-1273/Moderna N=7, and Ad26.COV2.S/Janssen N=10). Among these patients, only three (11%) patients were considered immunosuppressed per CDC criteria. Most breakthrough cases occurred when SAb-IgG was below the DCI laboratory designated antibody level for susceptibility of 2.0 U/L (n=23; 85%), with 18 (67%) of these cases occurring among individuals with SAb-IgG level below the assay manufacturer minimum detectable level of 1.0 U/L. Among the 19 individuals with 2 or more 2 SAb-IgG assessments prior to breakthrough infection, nine (47%) individuals always had SAb-IgG levels < 1 U/L consistent with vaccine nonresponse, 8 (42%) had waning SAb-IgG levels, and 2 (11%) consistently had maximum assay level. (**Figure panels A-C**).

## Discussion

SARS-CoV-2 vaccines are highly effective in maintenance dialysis patients with lower risk for COVID-19 cases, COVID-19 related hospitalization and, possibly, COVID-19 related death. Importantly, 26% of new infections and 27% of COVID related hospitalizations during the study period occurred among fully vaccinated patients reflecting that individuals receiving maintenance dialysis have compromised immune response.^9^

Recognizing this, several CDC vaccine recommendations highlight that maintenance dialysis patients are at high risk and may need additional vaccine doses. The timing of this additional dose, whether a third dose or a booster, remains uncertain. The need for more aggressive vaccine administration in dialysis has been well-demonstrated with vaccines versus hepatitis B vaccination.^10^ Nevertheless, the CDC did not specifically designate dialysis patients as immunocompromised persons who should receive routine administration of a third COVID-19 mRNA vaccine dose,^11^ limiting their definition to patients with primary immunodeficiency, advanced/untreated human immunodeficiency virus, receiving active cancer treatment, organ or stem cell transplant recipients and/or taking immunosuppressive medications. Other factors associated with low or no antibody response to SARS-CoV-2 vaccine not included in CDC criteria include older age, lower lymphocyte count, low serum albumin, higher body mass index, and prior non-response to hepatitis B vaccine, all of which are common among dialysis patients.^5,6,9^ We previously reported that 10-15% of dialysis patients were non-responders to 2 doses of mRNA vaccines.^3,12^ Recognizing this vulnerability, France authorized a third dose of SARS-CoV-2 mRNA vaccine for dialysis patients,^13^ reporting that a third dose of BNT162b2/Pfizer vaccine promoted seroconversion in more than half of initially non-responding dialysis patients.^14^

Breakthrough COVID-19 cases and COVID-19 related hospitalizations among fully vaccinated dialysis patients increased when delta variant became dominant, potentially superimposing a more virulent strain on waning immunity, as most dialysis patients were vaccinated March or earlier.^12^ Vaccine type may be associated with breakthrough infections, with receipt of the Ad26.COV2.S/Janssen vaccine not only associated with lower antibody response,^12^ but also with higher breakthrough and COVID-19 related hospitalization rates. Of note, the breakthrough COVID-19 case rate among fully vaccinated maintenance dialysis patients (1.6%) is higher than that reported for transplant patients (0.8%).^15^

Since IgG S1 antibody titers correlate with neutralizing titers and clinical efficacy,^16,17^ many clinicians associate detectable antibodies with clinical protection. The protective SAb-IgG titer threshold against COVID-19 infection has not been established. Of the 27 fully vaccinated patients with breakthrough infections who had antibody titers, the vast majority (85%) had very low or undetectable antibody levels prior to COVID-19 diagnosis; the majority never developed an initial antibody response to vaccine while most others waned to a very low level at the time of breakthrough COVID-19. We recently showed that more than half of fully vaccinated patients’ antibodies wane by 4-6 months, particularly among those whose initial response was not maximal based on our assay.^12^ The association of low or undetectable antibody levels with breakthrough COVID-19 supports potential use of antibody levels for guiding vaccination practices, and routinely monitoring COVID-19 vaccine seroresponse via SAb-IgG levels may identify patients at risk for COVID-19 infection. A similar model already exists for maintenance dialysis patients with Hepatitis B antibody monitoring.

Study strengths include the national population of a mid-size dialysis provider in the US with real world clinical outcomes. However, due to its observational design, residual biases may exit. Realistically, randomized clinical trials will not be performed in this population, accordingly observational data like these, viewed in the context of prior experience with vaccination for other conditions, are critical to guide clinical practice in the vulnerable maintenance dialysis population.

In conclusion, SARS-CoV-2 vaccines, particularly mRNA vaccines, are effective in maintenance dialysis patients, reducing the risk reduction of both COVID-19 cases and COVID-related hospitalization, albeit less than the general population.^18^ Current immunosuppression criteria are limited in identifying dialysis patients at highest breakthrough risk. Further research is needed to evaluate the potential utility of antibody titer monitoring to determine patients at highest risk for COVID-19 and the timing of additional vaccine administration.

## Data Availability

Data available upon request

## Figure Legend

Vaccine-Case number; M = mRNA-1273/Moderna; J = Ad26.COV2.S/Janssen; P = BNT162b2/Pfizer ★, immunocompromised per CDC criteria^11^

---

**Figure.**
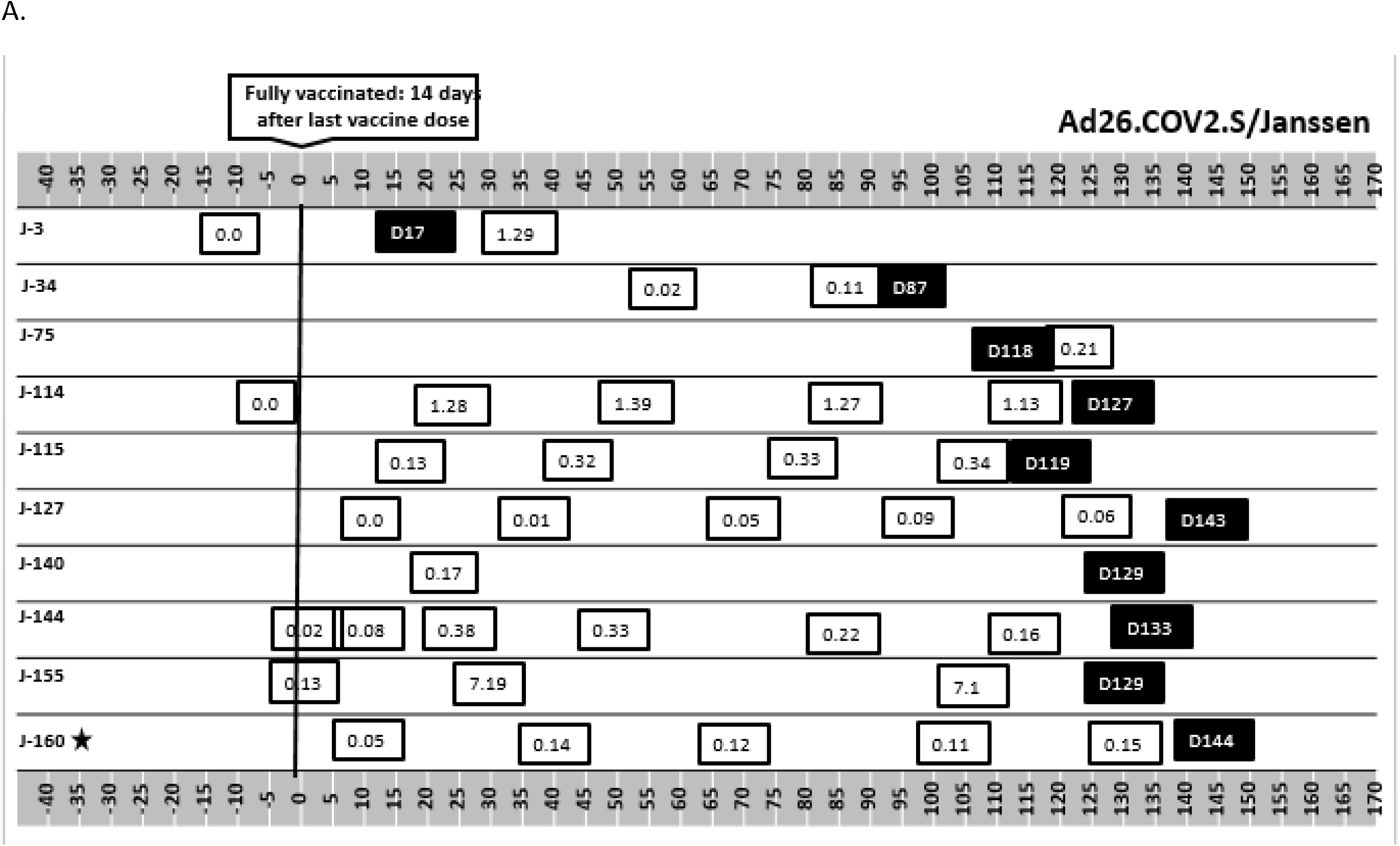
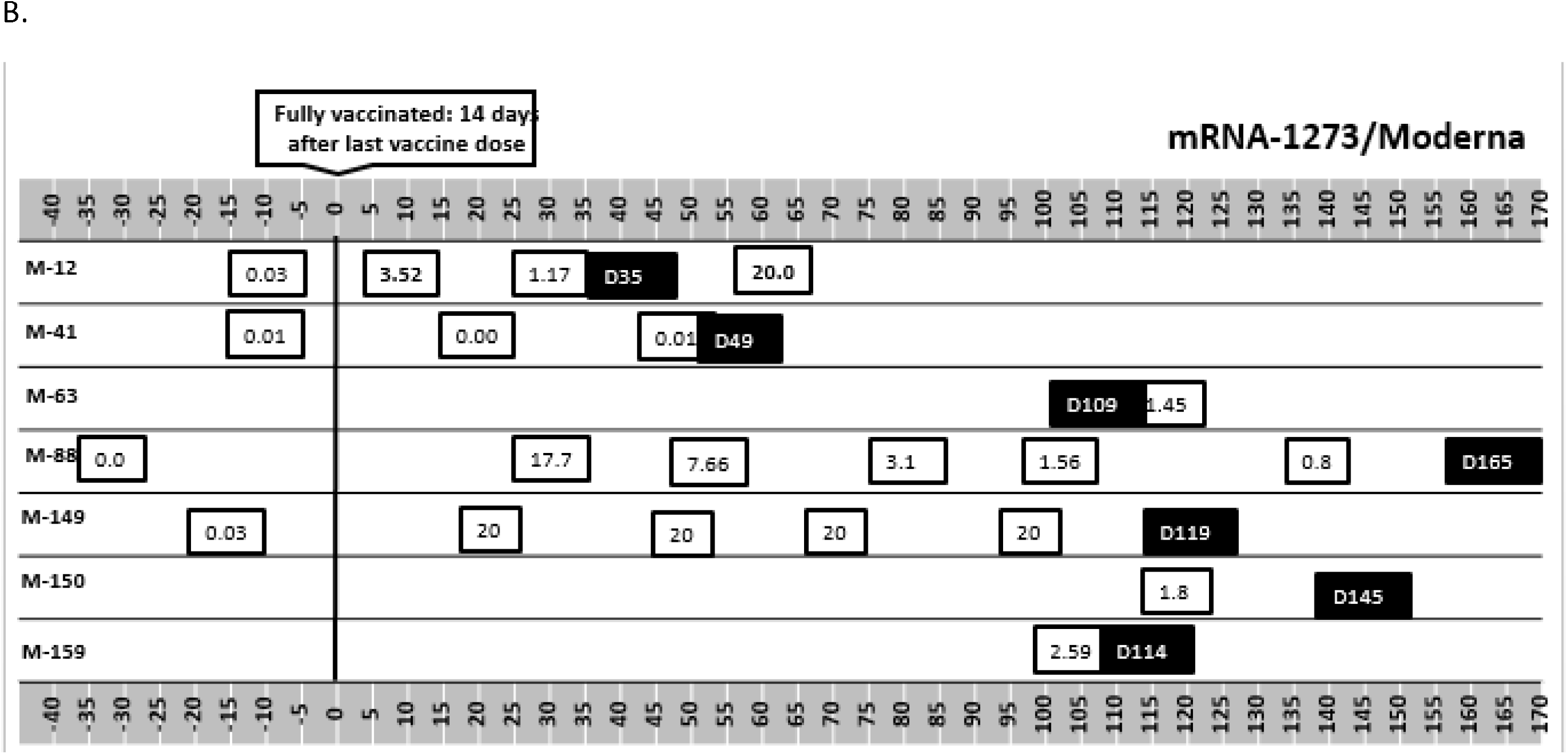
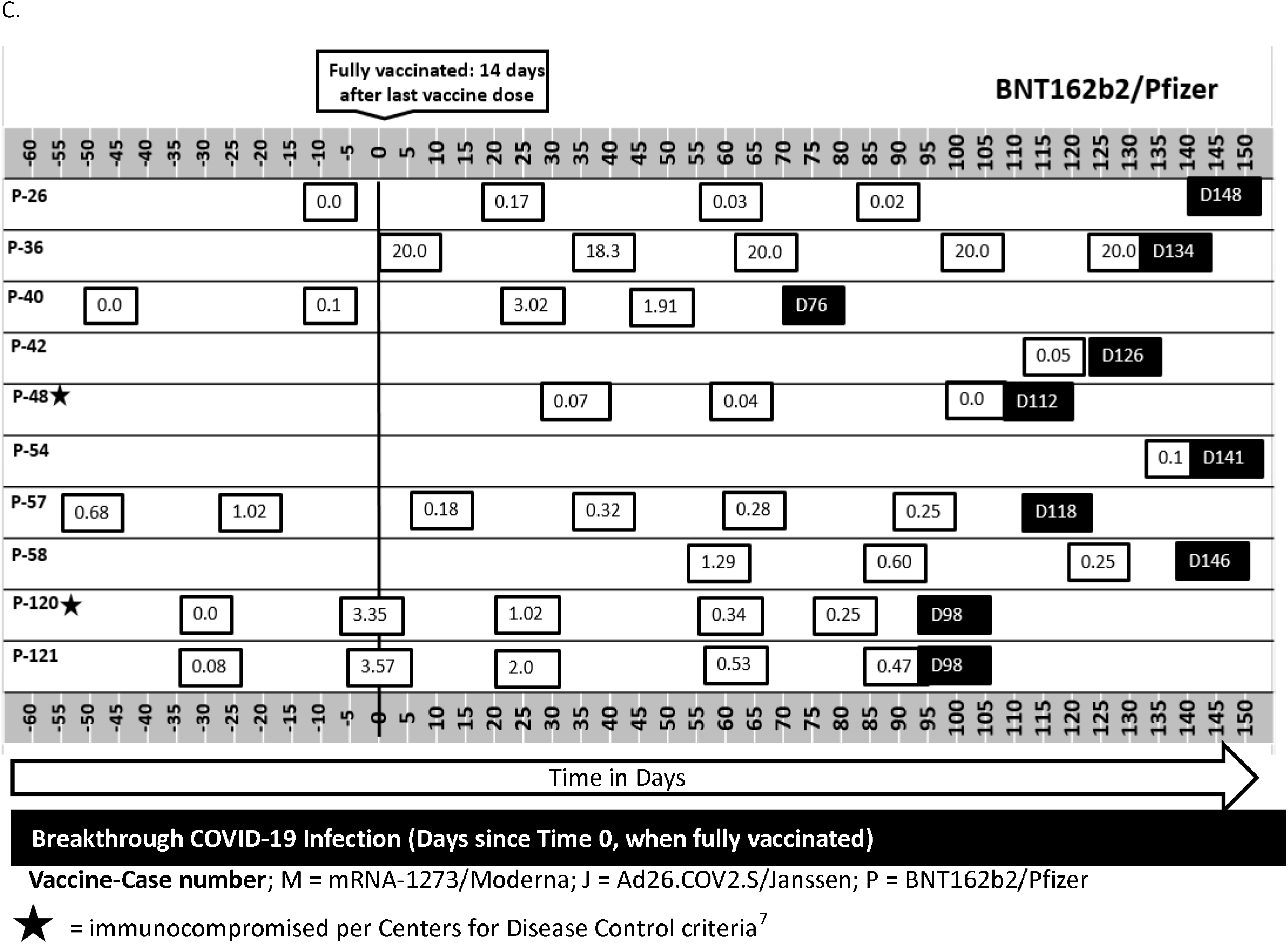
Patient SAb-IgG levels over time and relative to full vaccination status and COVID-19 infection

## Notes

### Competing Interest Statement

The authors have declared no competing interest.

### Funding Statement

no funding was obtained

### Author Declarations

This retrospective evaluation was reviewed and approved by WCG IRB (Work Order 1-1456342-1).

