## Supplement for "SARS-CoV-2 vaccine effectiveness and breakthrough infections in maintenance dialysis patients"

**Supplement Files**

Contents:


I. Detailed Methods
II. Table S1. Patient characteristics over entire study period (February 1-August 26, 2021
III. Table S2. New and breakthrough COVID-19 infections and COVID related hospitalizations pre delta variant predominant era
IV. Table S3. New and breakthrough COVID-19 infections and COVID related hospitalizations during delta variant dominate period.

### Methods

Study Population

Dialysis Clinic, Incorporated (DCI) is the largest non-profit provider of kidney care and dialysis care in the United States, operating 260 outpatient dialysis clinics in 29 states, 20 chronic kidney disease clinics in 5 states, and 3 organ procurement organizations in 3 states. All adult (age ≥18 years) maintenance dialysis patients treated between February 1 and August 27, 2021 in all DCI clinics contributed time-at-risk, with exceptions being any patient with known COVID-19 diagnosis prior to February 1, 2021, transient patients (e.g. visiting from a non-DCI clinic) and/or patients treated for acute kidney injury who were not designated as end-stage renal disease (ESRD).

Screening for COVID-19

All maintenance dialysis patients treated at DCI outpatient facilities, including home dialysis and in-center dialysis patients, are routinely screened for COVID-19 symptoms and any known exposure to infected person(s) at each clinic encounter. These symptoms include fever (≥100 degrees), sore throat, new or worsening cough or shortness of breath and loss of taste or smell with no other explanation. The vast majority of maintenance dialysis patients (>85%) are treated by in-center dialysis and are screened before each treatment, most often thrice weekly, while home dialysis patients are screened on average once each month. Exposure is defined by cumulative contact with a SARS-CoV-2 infected person during potential period of transmission, occurring within 6 feet and lasting for at least 15 minutes during a 24 hour period. Patients who screen positive are tested for SARS-CoV-2 either locally or through the DCI laboratory.

Most dialysis patients have multiple comorbid illnesses and have frequent contact with the healthcare system where they are tested even when asymptomatic. Similarly, many patients who are residents of group home settings, such as nursing homes, are frequently screened and tested for exposures as well. The diagnosis of COVID-19 is entered into DCI's electronic health record (EHR), often based on a report of a positive SARS-CoV-2 test result, which if performed via the DCI laboratory, are from nasal swabs tested by reverse transcriptase–polymerase chain reaction (RT-PCR) Cobas SARS-CoV-2 Assay [Roche Diagnostics].

SARS-CoV-2 Antibody Testing

The DCI central laboratory tests for serum IgG against the receptor binding domain of the S1 subunit of SARS-CoV-2 spike antigen using a US-FDA-EUA-approved chemiluminescent assay (ADVIA Centaur® XP/XPT COV2G). This semi-quantitative assay has a range between 0 and ≥20 U/L, with a "positive" result defined by a DCI-validated laboratory threshold of ≥2 U/L. The manufacturer defined threshold for a minimum antibody detection level is ≥1 U/L.[1] This test is available for physicians to order either one time or as part of a clinical testing protocol, but is not measured in all maintenance dialysis patients routinely. The clinical testing protocol follows monthly antibody titers from residual blood from routine monthly lab draws until the level is <1 U/L for two consecutive months or for up to 12 consecutive months.

Exposure and Time at Risk

Each eligible patient who received treatment for at least one day during the study period contributed days-at-risk. Patients could move from being unvaccinated (defined by the CDC to include the period extending up to 13 days after the first vaccine dose)[2] to partially vaccinated (for mRNA vaccines only; defined by the CDC as the period starting 14 days after the first mRNA vaccine dose up to 13 days after the 2nd mRNA vaccine dose) to fully vaccinated (≥14 days after completing the manufacturer recommended final dose). Patients could progressively contribute days-at-risk for each applicable category as long as they remained SARS-CoV-2 uninfected.

Outcomes

New COVID-19 diagnoses were documented based on screening as described above, or from contacts with other healthcare settings, such as hospitals and emergency department visits or clinic visits. All new COVID-19 diagnoses that occurred during the study period among eligible patients were each assigned to the appropriate vaccination status. For all breakthrough cases, the clinic was contacted directly to verify the reason why the patient was tested for SARS-CoV-2: for exposure to a known COVID-19 infected person or as required by a non-dialysis clinic/hospital protocol prior to providing a COVID-unrelated service/procedure (e.g. vascular access surgery) or as part of protocolized screening (e.g. nursing home). COVID-19 related hospitalizations were defined by documented primary diagnosis for the episode of care as COVID-19 (ICD-10 code U01.7).

Data Extraction and Analysis

All patient demographic and clinical variables used in this analyses were retrospectively obtained from the DCI EHR. These variables include: SARS-CoV-2 vaccine name and dates administered, age, sex (male or female), race (Black, White, Native American, Asian/Pacific Islander, Other/Unknown), ethnicity (Hispanic or Non-Hispanic), modality (in-center hemodialysis, home hemodialysis, peritoneal dialysis), date of ESKD, long term care facility status, body mass index, dialysis dose delivered (Kt/V), serum albumin, other vaccine use within 14 days of a COVID vaccine, hepatitis B surface antibody, immunosuppression markers (immune-modulating medications, prior transplant, immunodeficiency disorder), hospitalization within 14 days prior to vaccination, substance abuse disorder (alcohol or drug), and other comorbid conditions. For this study, any SARS-CoV-2 infection was considered a breakthrough COVID-19 case even if asymptomatic. Data integrity checks for COVID-19 documentation are performed weekly. These include comparing consistency of all available concurrent documentation, including: *de novo* ICD-10 COVID-19 diagnoses within the problem list, Patient Under Investigation status during symptom screening every visit, new COVID-19 diagnosis screening indicator during every visit, as well as any lab report indicating a positive test for SARS-CoV-2. Inconsistencies detected are referred to corporate nurses who directly communicated with individual clinic staff for resolution.

Case rates per 10,000 days at-risk for each vaccine status were compared using logistic regression with fully vaccinated patients as the reference group. Breakthrough cases among fully vaccinated patients were investigated for confirmation of COVID-19 infection or presence of immunocompromise (e.g., medications or medical conditions) at time of vaccination or COVID-19 case diagnosis. Odds ratio (OR) and 95% confidence intervals (CI) for COVID-19 case diagnosis and COVID-19 associated hospitalization rates per 10,000 days at-risk for each vaccine status (reference group full vaccination) and vaccine type (reference group mRNA-1273/Moderna) were compared using logistic regression and subsequently adjusted for all baseline patient characteristics that differed at baseline among vaccination status within each time period (Table S1). COVID-19 case and hospitalization rates were determined throughout study period, and further subdivided into pre-COVID-19 delta variant (February 1 - June 26, 2021) and COVID-19 delta variant predominance periods (June 27-August 27, 2021). Finally, a search was conducted to find any antibody titer results recorded in the EHR for patients who had breakthrough cases. These were arranged in a timeline where the dates when the patient was fully vaccinated (baseline) and the subsequent date of breakthrough infection relative to baseline were both represented. Results were de-identified and aggregated. This retrospective evaluation was reviewed and approved by WCG IRB (Work Order 1-1456342-1). Statistical analyses were performed using SAS v9.4.Reference:


1. COV2G, ADVIA Centaur XP and ADVIA Centaur CPT Systems. Published online July 2020. Accessed September 17, 2021. https://www.fda.gov/media/140704/download
2. Centers for Disease Control. Interim Clinical Considerations for Use of COVID-19 Vaccines Currently Approved or Authorized in the United States. Last accessed September 18, 2021. https://www.cdc.gov/vaccines/covid-19/clinical-considerations/covid-19-vaccines-us.html

Table S1. Patient characteristics over entire study period (February 1-August 26, 2021)

| | All Patients (N=13,223) | Fully (N=9,974) | Partial (N=441) | Unvaccinated (N=2,808) | P-value |
|---|---|---|---|---|---|
| New COVID-19 n (%) | 650 (4.3) | 168 (1.6) | 77 (11.5) | 405 (10.1) | <0.0001 |
| Time at Risk Median (IQR) | 207 (0) | 207 (0) | 207 (106) | 207 (29) | <0.0001 |
| Age (years) | 62.7 ± 14.8 | 64.7 ± 13.6 | 61.2 ± 14.9 | 57.9 ± 16.3 | <0.0001 |
| Age (≥65) | 7,339 (48.1) | 5,585(52.8) | 302 (45.0) | 1,452 (36.3) | <0.0001 |
| Age(Decade) | | | | | <0.0001 |
| <55 | 4,195 (27.5) | 2,365 (22.4) | 206 (30.7) | 1,624 (40.6) | |
| 55-64 | 3,717 (24.4) | 2,626 (24.8) | 163 (24.3) | 928 (23.2) | |
| 65-74 | 4,158 (27.3) | 3,118 (29.5) | 190 (28.3) | 850 (21.2) | |
| 75+ | 3,181 (20.9) | 2,467 (23.3) | 112 (16.7) | 602 (15.0) | |
| Female | 6,457 (42.3) | 4,305 (40.7) | 295 (44.0) | 1,857 (46.4) | <0.0001 |
| Male | 8,794 (57.7) | 6,271 (59.3) | 376 (56.0) | 2,147 (53.6) | |
| Race | | | | | <0.0001 |
| White | 7,121 (46.7) | 4,745 (48.1) | 267 (41.5) | 1,696 (43.8) | |
| Black | 5,466 (35.8) | 3,408 (34.6) | 260 (40.4) | 1,588 (41.0) | |
| Native American | 349 (2.3) | 259 (2.6) | 17 (2.6) | 57 (1.5) | |
| Asian/Pacific Islander | 480 (3.1) | 359(3.6) | 21 (3.1) | 78 (2.0) | |
| Other/Unknown | 1,835 (12.0) | 1,090(11.1) | 78 (12.1) | 450 (11.6) | |
| Hispanic | 893 (5.9) | 640(6.1) | 37 (5.5) | 216 (5.4) | 0.30 |
| Vintage (months) | 46.5 ± 55.9 | 46.8 ± 56.0 | 40.9 ± 52.7 | 42.7 ± 56.2 | <0.0001 |
| Body Mass Index (kg/m$^2$) | 28.6 ± 7.6 | 28.7 ± 7.4 | 28.3 ± 7.7 | 28.5 ± 8.2 | 0.11 |
| Long Term Care Facility | 1,731 (11.4) | 1,181 (11.2) | 108(16.1) | 442 (11.0) | 0.0004 |
| Home Dialysis | 2006 (13.2) | 1,424 (13.5) | 76(11.3) | 506 (12.6) | 0.15 |
| PD | 1,844 (12.1) | 1,299 (12.3) | 74(11.0) | 471 (11.8) | |
| HHD | 162 (1.1) | 125 (1.2) | 2(0.3) | 35 (0.9) | |

| | | | | | |
|---|---|---|---|---|---|
| Adequate Dialysis Dose | 11,313 (74.2) | 8,195 (77.5) | 497 (74.1) | 2,621 (65.5) | <0.0001 |
| Serum Albumin (g/dl) | 3.8 ± 0.5 | 3.8 ± 0.4 | 3.7 ± 0.5 | 3.7 ± 0.5 | <0.0001 |
| Other Vaccines within 14 days | 1,032 (6.8) | 959 (9.1) | 55 (8.2) | 18 (0.5) | <0.0001 |
| Pneumococcal | 262 (1.7) | 228 (2.2) | 25 (3.7) | 9 (0.2) | <0.0001 |
| Hepatitis B | 798 (5.2) | 755 (7.1) | 31 (4.6) | 12 (0.3) | <0.0001 |
| Influenza Vaccine | 47 (0.3) | 47 (0.4) | 0 (0.0) | 0 (0.0) | 0.99 |
| Hepatitis B seroimmunity[b] | 6,792 (44.5) | 6,426 (60.8) | 270 (40.2) | 96 (2.4) | 0.03 |
| Potential Immunosuppression | 2,470 (16.2) | 1,905 (18.0) | 122 (18.2) | 443 (11.1) | <0.0001 |
| Immune-modulating Medications | 1,509 (9.9) | 1,381 (13.1) | 94 (14.0) | 34 (0.9) | <0.0001 |
| Prior Transplant | 625 (4.1) | 583(5.5) | 25(3.7) | 17 (0.4) | 0.01 |
| Immunodeficiency Disorder | 1,271 (8.3) | 1,167(11.0) | 73(10.9) | 31 (0.8) | <0.0001 |
| Hospitalization within 14 days | 1,515 (9.9) | 1,314(12.4) | 140(20.4) | 61 (1.5) | <0.0001 |
| Disability | 695 (4.6) | 472 (4.5) | 31 (4.6) | 192 (4.8) | 0.69 |
| Tobacco Use | 2,456 (16.1) | 1,684 (15.9) | 123 (18.3) | 649 (16.2) | 0.25 |
| Alcohol Abuse Disorder | 1,538 (10.1) | 1,104 (10.4) | 73 (10.9) | 361 (9.0) | 0.03 |
| Drug Abuse Disorder | 777 (5.1) | 504 (4.8) | 41(6.1) | 232 (5.8) | 0.02 |
| Number of Comorbidities | 3.0 ± 1.8 | 3.0 ± 1.8 | 3.1 ± 1.8 | 2.8 ± 1.8 | <0.0001 |
| Diabetes Mellitus | 8,896 (58.3) | 6,326 (59.8) | 406 (60.5) | 2,164 (54.1) | <0.0001 |
| Hypertension | 12,504 (82.0) | 8,790 (83.1) | 562 (83.8) | 3,152 (78.7) | <0.0001 |
| Congestive Heart Failure | 3,357 (22.0) | 2,297 (21.7) | 161 (24.0) | 899 (22.5) | 0.28 |
| COPD[c] | 2,321 (15.2) | 1,638 (15.5) | 112 (16.7) | 571 (14.3) | 0.10 |
| Stroke/Cerebrovascular Disorder | 1,406 (9.2) | 1,002 (9.5) | 64 (9.5) | 340 (8.5) | 0.18 |

| | | | | | |
|---|---|---|---|---|---|
| Peripheral Vascular Disease | 1,981 (13.0) | 1,416 (13.4) | 101 (15.1) | 464 (11.6) | 0.004 |
| Thyroid Disorder | 2,322 (15.2) | 1,709 (16.2) | 84 (12.5) | 529 (13.2) | <0.0001 |
| History of Cancer | 1,461 (9.6) | 1,098 (10.4) | 52 (7.8) | 311 (7.8) | <0.0001 |

[a] Adequate dialysis defined by hemodialysis single pool Kt/V≥1.2 or peritoneal dialysis weekly Kt/V≥1.7.

[b] Hepatitis B seroimmunity defined as hepatitis B surface antibody ≥ 10 mIU/mL

[c] COPD Chronic Obstructive Pulmonary Disease

Patients are grouped based on their vaccination status at the end of follow-up. Unvaccinated includes patients who never received a vaccine or recipients of a single dose of a vaccine within 14 days of vaccine receipt; partially vaccinated includes patients who were ≥14 days after the first mRNA vaccine dose but <14 days after the 2nd mRNA vaccine dose; fully vaccinated patients includes all patients ≥14 days after the last vaccine dose. For the table, categories are mutually exclusive.

Table S2. New and breakthrough COVID-19 infections and COVID related hospitalizations during the pre delta variant predominant era

| Status/Category | Person Days-at risk | # Events | Events per 10,000 Patient Days | Odds Ratio (95% Confidence Interval) | |
|---|---|---|---|---|---|
| | | | | Unadjusted | Adjusted |
| **COVID Cases by Vaccination Status** | | | | | |
| **Unvaccinated* (N = 3,869)** | 931,411 | 292 | 3.14 | 9.32 (6.24,13.92) | 8.89 (5.92,13.34) |
| **Partially Vaccinated* (N = 643)** | 275,589 | 67 | 2.43 | 7.23 (4.60,11.36) | 7.10 (4.51,11.16) |
| **Fully Vaccinated* (N = 9,861)** | 773,479 | 26 | 0.34 | Reference | Reference |
| **COVID Cases by Vaccine Type** | | | | | |
| **Ad26.COV2.S/Janssen (N = 460)** | 33,513 | 2 | 0.60 | 2.01 (0.45,8.91) | 1.75 (0.38,8.01) |
| **BNT162b2/Pfizer (N = 3,839)** | 302,075 | 11 | 0.36 | 1.23 (0.55,2.74) | 1.07 (0.48,2.42) |
| **mRNA-1273/Moderna (N = 5,562)** | 437,891 | 13 | 0.30 | Reference | Reference |
| **COVID-related Hospitalization by Vaccination Status** | | | | | |
| **Unvaccinated (N = 3,869)** | 990,623 | 89 | 0.90 | 5.43 (3.03,9.72) | 5.20 (2.89,9.42) |
| **Partially Vaccinated (N = 643)** | 287,183 | 26 | 0.91 | 5.47 (2.81,10.65) | 5.33 (2.74,10.37) |
| **Fully Vaccinated (N = 9,861)** | 785,547 | 13 | 0.17 | Reference | Reference |
| **COVID-related Hospitalization by Vaccine Type** | | | | | |
| **Ad26.COV2.S/Janssen (N = 460)** | 34,199 | 2 | 0.58 | 6.51 (1.19,35.53) | 7.45 (1.25,44.52) |
| **BNT162b2/Pfizer (N = 3,839)** | 306,234 | 7 | 0.23 | 2.54 (0.75,8.69) | 2.42 (0.70,8.38) |
| **mRNA-1273/Moderna (N = 5,562)** | 445,114 | 4 | 0.09 | Reference | Reference |

* Although patients can contribute time to any vaccination status, the N in the first column refers to patients' status at the end of follow-up. Unvaccinated includes patients who never received a vaccine or recipients of a single dose of a vaccine within 14 days of vaccine receipt; partially vaccinated includes patients who were ≥14 days after the first mRNA vaccine dose but <14 days after the 2nd mRNA vaccine dose; fully vaccinated patients includes all patients ≥14 days after the last vaccine dose.

Table S3. New and breakthrough COVID-19 infections and COVID related hospitalizations during delta variant dominate period.

| Status/Category | Person Days-at risk | # Events | Events per 10,000 Patient Days | Odds Ratio (95% Confidence Interval) | |
|---|---|---|---|---|---|
| | | | | Unadjusted | Adjusted |
| **COVID Cases by Vaccination Status** | | | | | |
| **Unvaccinated* (N = 2,808)** | 176,127 | 110 | 6.25 | 2.58 (2.10,3.31) | 2.27 (1.72,3.00) |
| **Partially Vaccinated* (N = 441)** | 25,037 | 9 | 3.59 | 1.48 (076,2.91) | 1.28 (0.65,2.52) |
| **Fully Vaccinated* (N = 9,974)** | 586,244 | 142 | 2.42 | Reference | Reference |
| **COVID Cases by Vaccine Type** | | | | | |
| **Ad26.COV2.S/Janssen (N = 464)** | 27,126 | 13 | 4.79 | 2.68 (1.47,4.88) | 2.36 (1.29,4.32) |
| **BNT162b2/Pfizer (N = 3,934)** | 229,608 | 70 | 3.05 | 1.70 (1.20,2.41) | 1.68 (1.18,2.38) |
| **mRNA-1273/Moderna (N = 5,576)** | 329,510 | 59 | 1.79 | Reference | Reference |
| **COVID-related Hospitalization by Vaccination Status** | | | | | |
| **Unvaccinated (N = 2,808)** | 179,533 | 46 | 2.56 | 3.10 (2.07,4.64) | 3.00 (1.89,4.75) |
| **Partially Vaccinated (N = 441)** | 25,307 | 6 | 2.37 | 2.87 (1.23,6.70) | 2.48 (1.05,5.86) |
| **Fully Vaccinated (N = 9,974)** | 592,774 | 49 | 0.83 | Reference | Reference |
| **COVID-related Hospitalization by Vaccine Type** | | | | | |
| **Ad26.COV2.S/Janssen (N = 464)** | 27,578 | 4 | 1.45 | 3.02 (1.01,9.03) | 2.54 (0.84,7.67) |
| **BNT162b2/Pfizer (N = 3,934)** | 232,400 | 29 | 1.25 | 2.60 (1.41,4.78) | 2.58 (1.40,4.78) |
| **mRNA-1273/Moderna (N = 5,576)** | 332,796 | 16 | 0.48 | Reference | Reference |

* Although patients can contribute time to any vaccination status, the N in the first column refers to patients' status at the end of follow-up. Unvaccinated includes patients who never received a vaccine or recipients of a single dose of a vaccine within 14 days of vaccine receipt; partially vaccinated includes patients who were ≥14 days after the first mRNA vaccine dose but <14 days after the 2nd mRNA vaccine dose; fully vaccinated patients includes all patients ≥14 days after the last vaccine dose.
